# Efficacy of Tocilizumab in Covid 19: A metanalysis of case series studies

**DOI:** 10.1101/2020.08.12.20173682

**Authors:** Shalam Mohamed Hussain, Ayesha Farhana Syeda, Osama Al-Wutayd, Abdullah Al Nafeesah, Mohammad Alshammari, Sulaiman Alnasser, Nandakumar Krishnadas

**Affiliations:** Former faculty and independent reseaarcher, College of Pharmacy, Qassim University, Saudi Arabia; Department of Pharmaceutics, Unaizah College of Pharmacy, Qassim University; Department of Family and Community Medicine, Unaizah College of Medicine and Medical Sciences, Qassim University, Saudi Arabia; Department of Pediatrics, Unaizah College of Medicine and Medical Sciences, Qassim University, Saudi Arabia; Department of Pharmacy Practice, Unaizah College of Pharmacy, Qassim University; Department of Pharmacology, Unaizah College of Pharmacy, Qassim University; 6 Department of Pharmacology, Manipal College of Pharmaceutical Sciences, India

**Keywords:** COVID 19, tocilizumab, interleukin-6, cytokine storm, case series

## Abstract

**Background:** characteristic feature of COVID-19 during its progression in severity is the cytokine storm (via interleukin-6, IL-6) which is responsible for secondary acute respiratory distress syndrome (ARDS) [4]. Tocilizumab has an antagonist effect on the IL-6 receptor. With present review and metanalysis, we intend to update the current status on the clinical efficacy of tocilizumab in the treatment of Covid 19 infections in the published literature of case series.

**MATERIALS AND METHODS:** The following inclusion criteria were used: (i) case series studies (number of reported patients in each study equal to or greater than ten (ii) use of tocilizumab alone or in combination with standard of care therapy (iii) Covid 19 adult patients (iv) the studies with endpoints on all-cause mortality, need for mechanical ventilation, clinical improvements.

**Data synthesis and statistical analysis:** Meta-analysis was performed using a random effects model and the DerSimonian and Laird method.

**RESULTS:** 18 were selected for the quantitative analysis (meta-analysis). 14 studies were retrospective and 4 were prospective

**Meta-analysis:** The mortality rate of COVID-19 patients with tocilizumab was 21% (251/1212) Asymmetric funnel plot in the cylindrical form due to publication bias

**In conclusion:** the present synthesis provide us useful insights with the other available evidence to refine our strategy and equip ourselves effectively with tocilizumab to defeat COVID 19 to save humanity.

**Limitations:** The included studies utilized varied doses of tocilizumab (single or double), and duration drug availability issues emerged in some centers, which may have influenced both sample sizes and study designs.

**Clinical implications:** Incorporated studies without control groups into systematic reviews and quantitative synthesis especially when there are no other studies to consider can provide information for formulating effective treatment strategies for management of COVID 19 infections through the use of tocilizumab.

## 1. Introduction

Since December 2019, when in Wuhan, China, coronavirus disease 2019 (COVID-19) first identified, has spread rapidly in many countries seriously impacting the global economy[1]. Apart from the classically known symptoms of COVID-19 fever, cough, and shortness of breath several studies discovered additional clinical manifestation of the disease was such as fatigue, myalgia, increased septum production, chest pain, chill), headache, sore throat and diarrhea[2]. The Centers for Disease Control and Prevention (CDC) also added new symptoms including new loss of taste or smell, repeated shaking with chills, and sore throat. A characteristic feature of COVID-19 during its progression in severity is the cytokine storm (via interleukin-6, IL-6) which is responsible for secondary acute respiratory distress syndrome (ARDS)[2][3][4]. ARDS is the most devastating complication of SARSCoV-2 with a higher death toll[5][6]. The incidence of ARSD following COVID-19 is higher in severe cases. Hence treating cytokine storm has resulted in rescuing COVID-19 affected patients. Currently, there is no specific drug for COVID-19 induced cytokine storm but the tocilizumab is a recombinant humanized monoclonal antibody that has an antagonist effect on the IL-6 receptor. It is currently used in the treatment of rheumatoid arthritis, but could also play a key role in treatment for severely ill patients with COVID-19[7][8]. Tocilizumab has received considerable attention in studies finding reduction in COVID-19 related deaths and morbidities. As the treatment of cytokine storm induced by COVID-19 with tocilizumab has broad prospects, it is been recommended in many countries as well as been used off label[9][10]. However, the clinical experience and data of tocilizumab in the treatment of COVID-19 are limited, despite several reviews and metanalyses about its use in COVID 19. Though the randomized controlled trials (RCTs) provide rigorous results for effectiveness of any interventions, yet may not be the enough for all questions[11]. However, in this pandemic tiring times, difficult to complete and investigate the suitable remedies for COVID 19. It is also important to consider the best possible evidence in these difficult times to find solution for the devastating COVID 19.

Systematic reviews are an established approach to identifying and summarizing a body of literature associated with a particular topic area and meta-analysis (the formal statistical pooling of data from multiple studies), can be used to develop summary estimates for proportions from case series studies. Previous reviews and metanalyses on the utility of tocilizumab have explored on the available evidence of case control studies, a large body of evidence, such as case series, however, was left out. Case series studies can also provide valuable additional evidence for practitioners and decision makers especially in this hard time of war like situation against COVID 19 pandemic[12][13]. Currently, there is no standardized method for synthesizing results of studies that do not have control groups but several studies published with case series have provided valuable insight into the decision-making processes. With present review and metanalysis, we intend to update the current status on the clinical efficacy of tocilizumab in the treatment of Covid 19 infections in the published literature of case series.

## 2 MATERIALS AND METHODS

### 2.1 Literature search strategy

A preliminary search conducted prior to formal literature review found limited evidence on the topic and yet patients being treated with tocilizumab, revealing an urgency in dealing with this issue. Therefore, the present endeavor is an attempt to contribute to enlighten the success against this growing menace. A literature search was conducted on PubMed, Google Scholar and repositories of preprints (MedRxiv) for articles published until July, 2020 using keywords such as “COVID-19,” “tocilizumab,” “interleukin 6 antagonist,” or “IL-6 blocker”, or “human monoclonal antibody,” “cytokine storm treatment”). In Google scholar text word search of titles and abstracts was conducted using the following search terms ‘Tocilizumab,’ ‘anti-interleukin-6 antibody,’ and ‘COVID-19’ or ‘coronavirus 2019’ in various combinations. This meta-analysis was performed in accordance with preferred reporting items for systematic review and meta-analysis statement (PRISMA).

### 2.2 Inclusion and exclusion criteria

The following inclusion criteria were used: (i) case series studies (number of reported patients in each study equal to or greater than ten (ii) use of tocilizumab alone or in combination with standard of care therapy (iii) Covid 19 adult patients independently of the severity of their symptoms and (iv) the studies specified any or all of the following endpoints on all-cause mortality, need for mechanical ventilation, clinical improvements, ICU admissions, incidence of adverse outcomes, length of hospital stay (v) positive SARS-CoV-2 diagnosis by reverse-transcriptase polymerase chain reaction. The following outcomes were considered as clinical efficacy measurements: survival rate, resolution of symptoms and discharge from the hospital. No limits were applied to either date or language of the literatures published. Review articles, case reports with less than 10 cases, cases reporting pediatric or pregnant COVID 19 or organ transplanted patients were excluded from the quantitative analysis. The articles, which were in the form of only abstracts, letters without original data, meta-analysis, animal studies, or studies that did not report original data on tocilizumab were excluded. Further attempts were made to identify relevant studies through references to find eligible studies. The flow of information from identification to inclusion of studies is summarized in figure 1.

**Figure 1.**
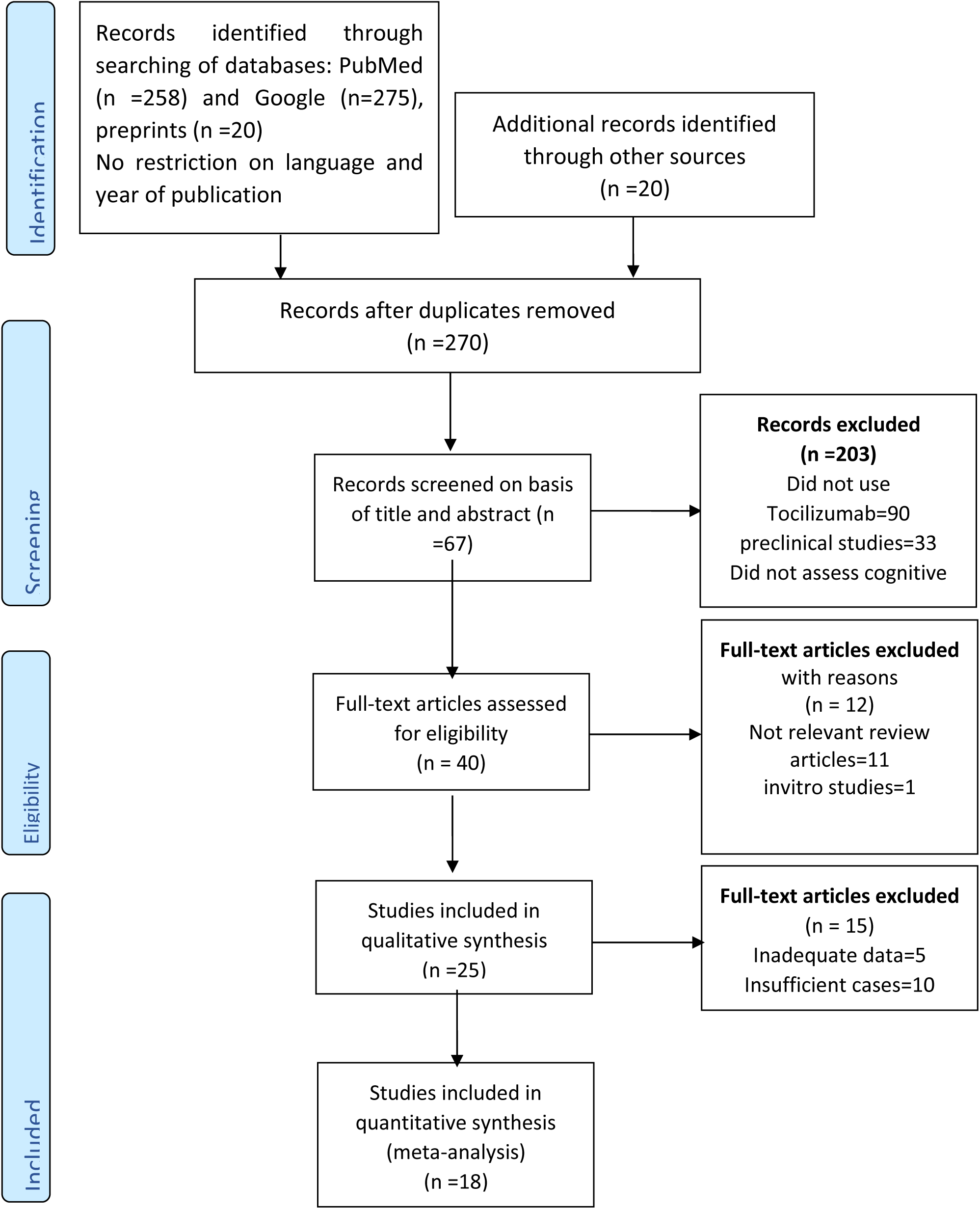
Flow chart of study selection process. The number of studies in each phase is indicated between brackets.

### 2.4 Data extraction

A standard data extraction template was used by two authors independently to extract the following information from each included study: (1) article information (author and publication year, country, type of study, number of patients in tocilizumab groups, patients’ gender, age, drugs used in standard of care, and mortality in both groups. (3) type of intervention; type, dose, duration, and frequency of administration of tocilizumab (4) type of outcome measure, mortality in severe COVID-19 patients who received tocilizumab, improvement proportions, number for mechanical intervention or ICU admissions. Secondary outcomes were proportions reporting any adverse events and time in days to recover or discharge. The characteristics of the studies extracted are shown in Table 1. The proportions of individuals meeting each outcome will be included in meta-analysis to provide summary estimates of proportions. Eligibility assessment was performed independently in an unblinded standardized manner by two reviewers. Disagreements between reviewers, if any, were resolved by consensus.

**Table 1.** Case series examining the effect of Tocilizumab COVID 19 patients.

| **S.No** | **Author, year** | **Country** | **Type of study<sup>#</sup>** | **Population (n)** | **Age in yrs**<br>**Mean**<br>**(range)** | **Male**<br>**(n)** | **Common comorbidities<sup>#</sup>** | **TCZ doses, duration** | **Standard of care<sup>#</sup>** | **Adverse events** |
| --- | --- | --- | --- | --- | --- | --- | --- | --- | --- | --- |
| **1** | **Conrozie** | **France** | **R** | **40** | **73.7 (65-89)** |  | **HT, DM, COPD or emphysema** | **8mg/kg/day, IV** | **Amoxiclav/ceft/lev/LMWH** | **NR** |
| **2** | **Marfella** | **Italy** | **R** | **78** | **NR** |  | **DM** | **8 mg/kg/12 h, IV, two doses** | **NR** | **NR** |
| **3** | **Patel** | **India** | **R** | **20** | **54 (47-63)** | **17** | **DM, HT, IHD** | **8 mg/kg/12 h, IV, two doses** | **HCQ/AZT/ceft/LMWH** |  |
| **4** | **Betul Borku** | **Turkey** | **P** | **12** | **65.83(47-69)** | **6** | **DM, HT, COPD, malignancy, chronic renal failure** | **IV 400 mg, two doses in 24hrs** | **HCQ/AZT/oseltamivir/moxifloxacin** | **NR** |
| 5 | Paola, | Italy | P | 100 | 62<br>(57–71) | 88 | DM,<br>cardiovascular disease,<br>COPD,<br>chronic renal failure | 8 mg/kg/12 h, IV, two doses | NR | NR |
| 6 | John Knorr | USA | R | 66 | age ≥ 18 | | DM, HT, asthma or COPD | 8mg/kg, 3 doses/24hrs | HCQ | NR |
| 7 | Pan Luo | china | R | 15 | 73 (62-80) | 12 | CVD and endocrine diseases | 80 to 600 mg per time | Glucocorticoids | NR |
| 8 | Xiaoling Xua | china | R | 21 | 56.8 (25-88) |  | HT, DM, COPD, CHD | 8mg/kg, two doses/24hrs | Lopinavir/ritonavir//IFN | NR |
| 9 | Christina | USA | R | 153 | 64 (22-99) | 125 | DM, immunosuppressed, COPD, obesity | 8mg/kg, two doses/24hrs | HCQ/Glucocorticoids/antivirals | NR |
| 10 | Andrew | USA | R | 134 | 62 (53 - 70) | 99 | DM, COPD, HT, Renal failure, obesity | 400mg, single dose | None | NR |
| 11 | Shoji Hashimoto | JAPAN | R | 13 | 61.6 ± 14.7 | 11 | DM, HT, COPD, Dyslipidemia, RA | 400mg, single dose | Ciclesonide/Favipiravir/Methylprednisolone | NR |
| 12 | Rand Alattar | Qatar | R | 25 | 58(50-63) | 23 | DM, chronic kidney disease, and CVD | median total dose of 5.7 mg/kg | HCQ/AZT/lopinavir/ritonavir/INT | Anemia, rise in ALT and QT interval prolongation |
| 13 | Sciascia | ITALY | P | 63 | 62.6±12.5 | 56 | DM, chronic kidney disease, and CVD | 8 mg/kg, IV two doses | Lopinavir/ritonavir/<br>Darunavir/cobicistat | NR |
| 14 | Valentina | ITALY | P | 51 | 60 (50-70) | 40 | CVD, HT,<br>DM,<br>Chronic lung<br>diseases,<br>Cancer, RA | 8 mg/kg, IV<br>two doses | NR | ↑ hepatic<br>enzymes,<br>thrombocy<br>topenia,<br>and<br>serious<br>bacterial<br>and fungal<br>infections |
| 15 | Russell | USA | R | 145 | 58.1 | 93 | DM, COPD,<br>chronic<br>cardiac or<br>renal disease,<br>immunodefici<br>ency | 8 mg/kg, IV<br>ONE TO<br>two<br>doses/24 h | NR | NONE |
| 16 | Miguel | SPAIN | R | 186 | 65 | 126 | DM, COPD,<br>Vasculopathy,<br>Immunosuppr<br>ession |  | Lopinavir/riton<br>avir/HCQ/DOX/<br>AZT/LMWH | ↑ hepatic<br>enzymes,<br>creatinine,<br>hyperkale<br>mia |
| 17 | Daria | RUSSIA | R | 89 | 53 |  | DM, COPD, |  | NR | NR |
| 18 | Casey A.<br>Rimland | USA | R | 11 | 59<br>(48-65) | 9 | HT, DM,<br>Chronic lung<br>diseases,<br>Obesity | 8 mg/kg, IV<br>one to two<br>doses/24 h | NR | ↑ liver<br>enzymes |

Abbreviations used in the table above, R: retrospective study, P: retrospective study, NR: Not Reported, HT: hypertension, DM: diabetes mellitus, COPD: chronic pulmonary disorder, Amoxiclav: amoxycillin and clavulanic acid combination, ceft: ceftriaxone, lev: levofloxacin, LMWH: low molecular weight heparins, INF: interferons, AZT: azithromycin, ALT: alanine aminotransferase, RA: rheumatoid arthritis

### 2.4 Quality assessment of the included studies

The quality of all studies was assessed independently by two reviewers using the “Quality Appraisal Checklist for Case Studies” tool developed by the Institute of Health Economics [11]

### 2.5 Data synthesis and statistical analysis

The primary outcome was the proportion of patients with COVID 19 who were being treated with tocilizumab. This was calculated as the number of patients being affected by the total number of cases of COVID 19. Standard errors and confidence intervals for a single proportion were derived. The derived proportions were adjusted by logit transformation. Meta-analysis was performed using a random effects model and the DerSimonian and Laird method. Random effect models and I-square (I^2^) test were used to assess quantitatively the impact of anticipated study heterogeneity on the results of the meta-analysis. p value <.1 and I^2^ + value of >50% were considered statistically significant. In proportional meta-analysis, a common challenge, that is faced is to deal with proportions when they are too large or too small (close to 0 or to 1) which causes the variance of the proportion to be very small leading to an inappropriate large weight. To over this, a count of 0.5 was added to or subtracted from the number of COVID 19 cases to those reporting an outcome of 0% or 100% respectively and to conduct the meta-analysis by transforming the proportions using logit transformation[14][15]. Forest plot was generated to depict the logit proportion along with its 95% confidence interval for each study as well as the pooled them.

## 3 RESULTS

### 3.1 Study selection

The electronic search strategy retrieved from PubMed (n =258) and Google (n=275), preprints (n =20) and other sources (n =20). From this 270 were left after removing duplicates. Following screening of title and abstracts (n =67) were retained for full text article assessment. Forty papers met our inclusion criteria. Further, 25 papers reporting the investigation of efficacy of tocilizumab on COVID 19 were included in the qualitative analysis and finally eighteen studies were selected for the quantitative analysis (meta-analysis).

### 3.2 Study characteristics

The characteristics of the included studies are summarized in Table 1. There was a large variation in the characteristics of selected studies due to relatively short course of disease (7 to 35 days) and were considered to have not a satisfactory follow up duration. 14 studies were retrospective and 4 were prospective nonrandomized case series. Eighteen studies [9], [16], [24]-[33], [17], [34], [18]-[20], [20]-[23] that assessed mortality, 13 also reported clinical improvements in COVID 19 patients and 15 assessed the need for mechanical ventilation by the patients. Among countries where case series reported, USA-5, Italy-4, China-2, France-1, Qatar-1, India-1, Turkey-1, Japan-1 Spain=1 and Russia-1. Common co-morbidities reported in COVID 19 patients were hypertension, diabetes mellitus, chronic pulmonary disorder, obesity, rheumatoid arthritis and kidney disease. Only few studies treating COVID 19 patients with tocilizumab did not report any adverse events. The reported common events were ↑ hepatic enzymes, creatinine, hyperkalemia, serious bacterial and fungal infections, anemia, rise in ALT and QT interval prolongation. Majority of the studies adopted standard of care treatment (SOC) along with one to three doses of tocilizumab. The commonly employed drugs as part of SOC were amoxycillin and clavulanic acid combination, ceftriaxone, levofloxacillin, low molecular weight heparins, interferons, azithromycin. The standard recommended regimen for tocilizumab is is 4 to 8 mg/kg to a maximum of 800 mg per dose, with an additional dose 8 to 12 hours later if clinically required. All the included studies found to follow this regimen (Table 1).

### 3.3 Quality assessment of the included studies

Out of eighteen studies, 14 were retrospective ref and 4 studies were prospective nonrandomized case series. The quality of each study was assessed according to the Institute of Health Economics Quality Appraisal Checklist for Case Series Studies[11] is provided in Table 2. Overall scores based on the positive attributes in varied from 9to 18 out of a possible 20. The study conducted by Sciascia et al had the highest score, with 18 points, whereas study by Rand et al had a score of 17. Studies by Andrew et al, Shoji et al and Betul Borku et al had a score of 16. Score for other studies ranged from 9 to 15. The overall lowest score (5) was for the questions, description of co-interventions and prospective type of study. Therefore, most studies were of poor to moderate quality according to the checklist.

**Table 2:**
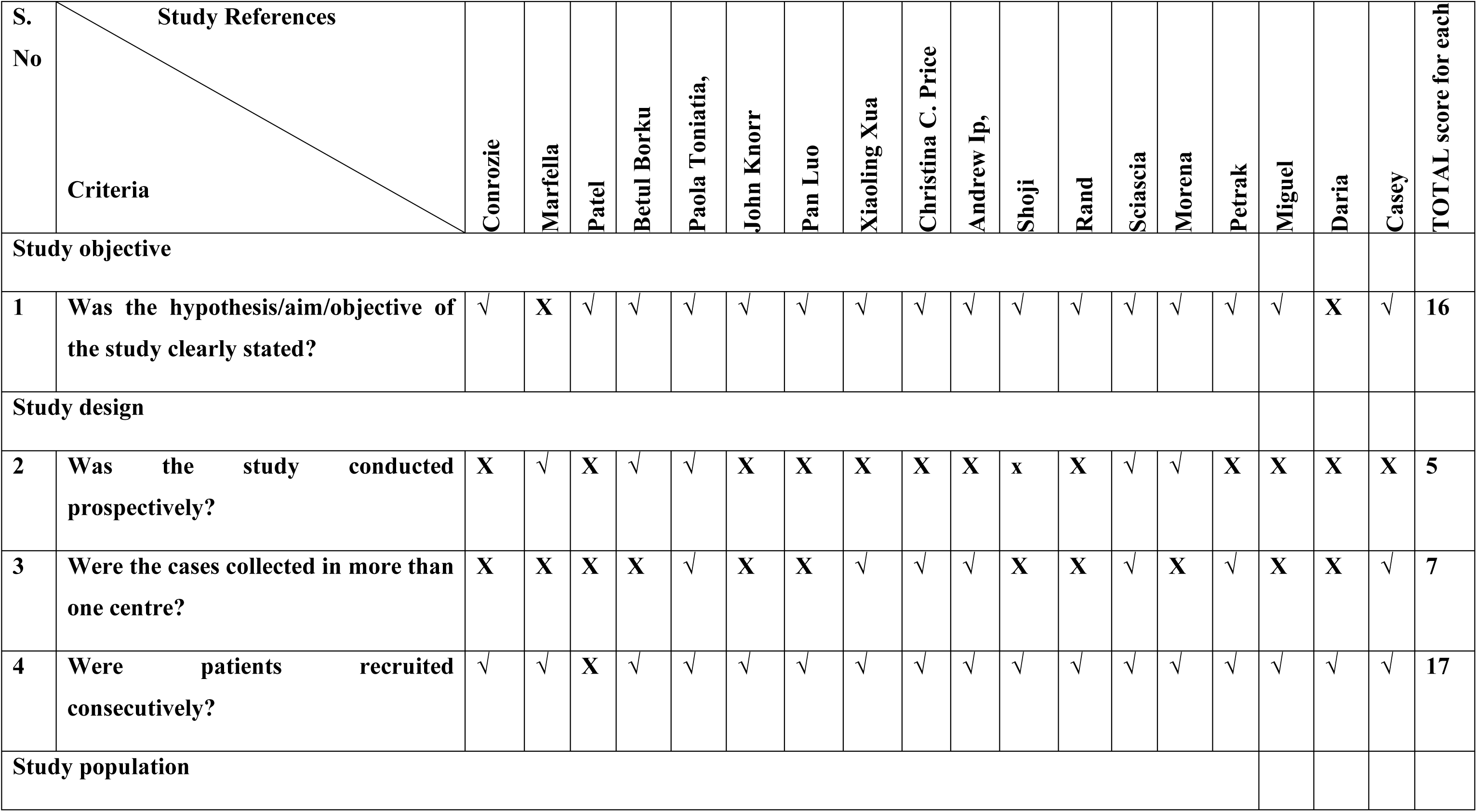
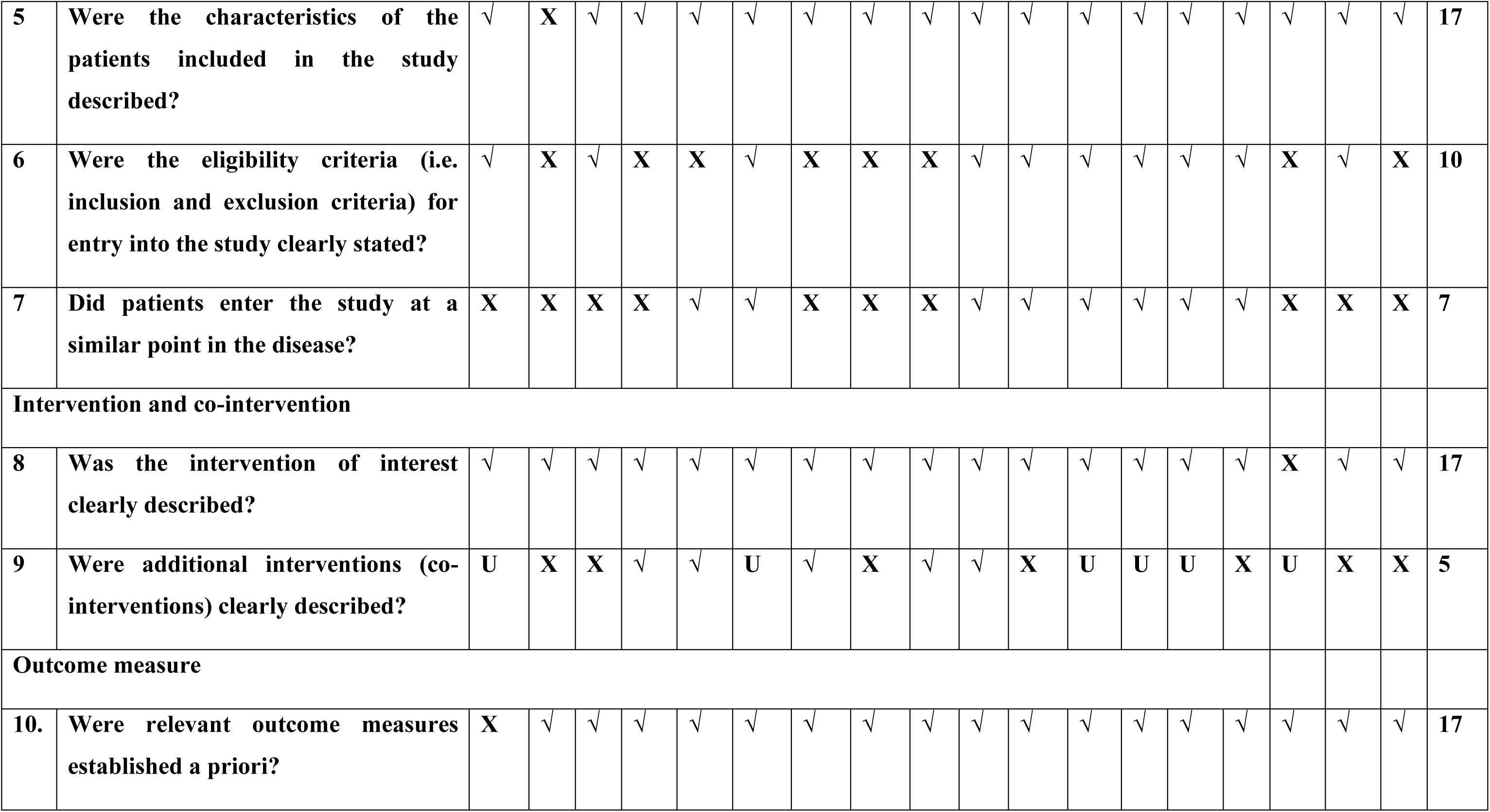
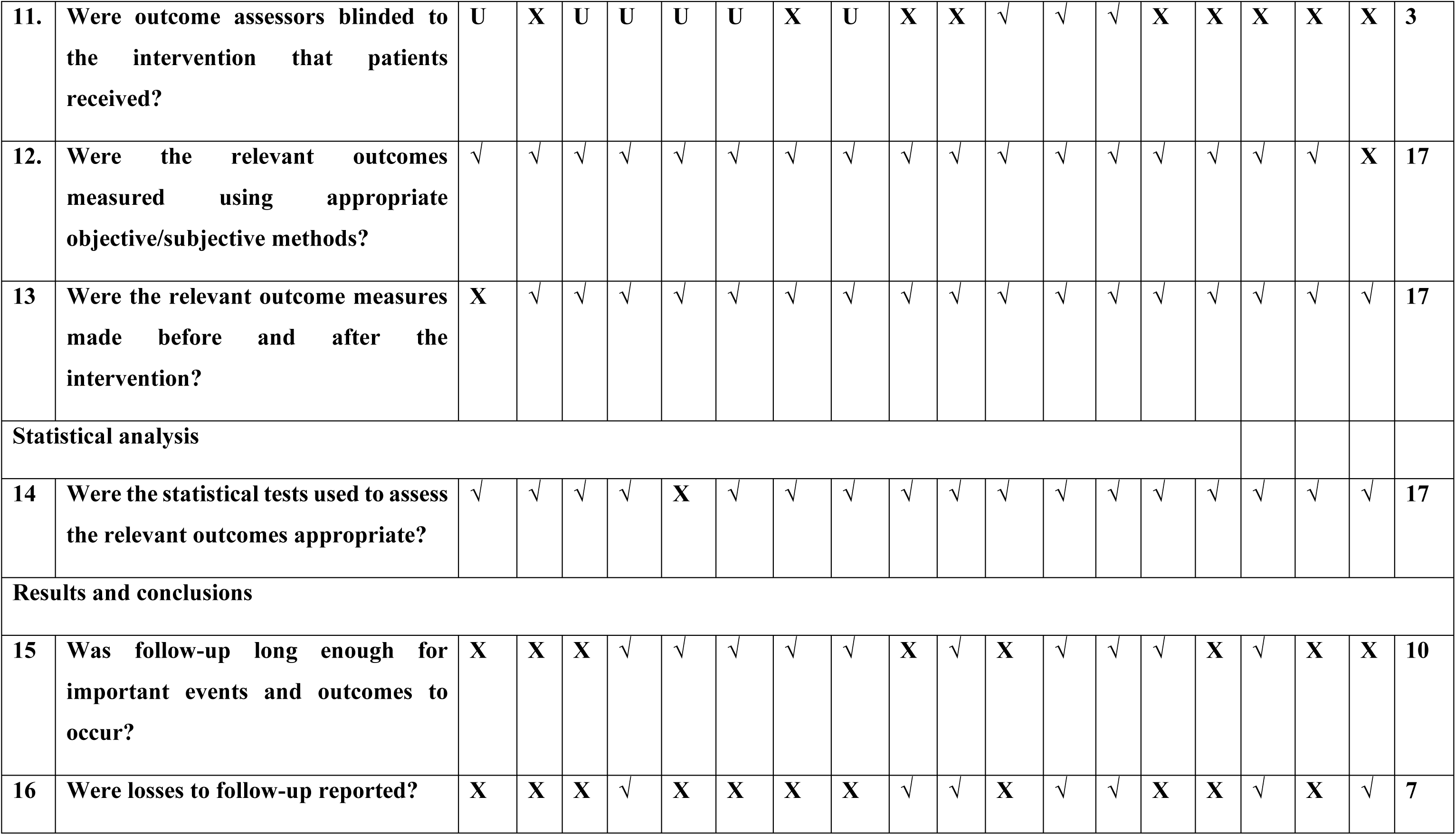
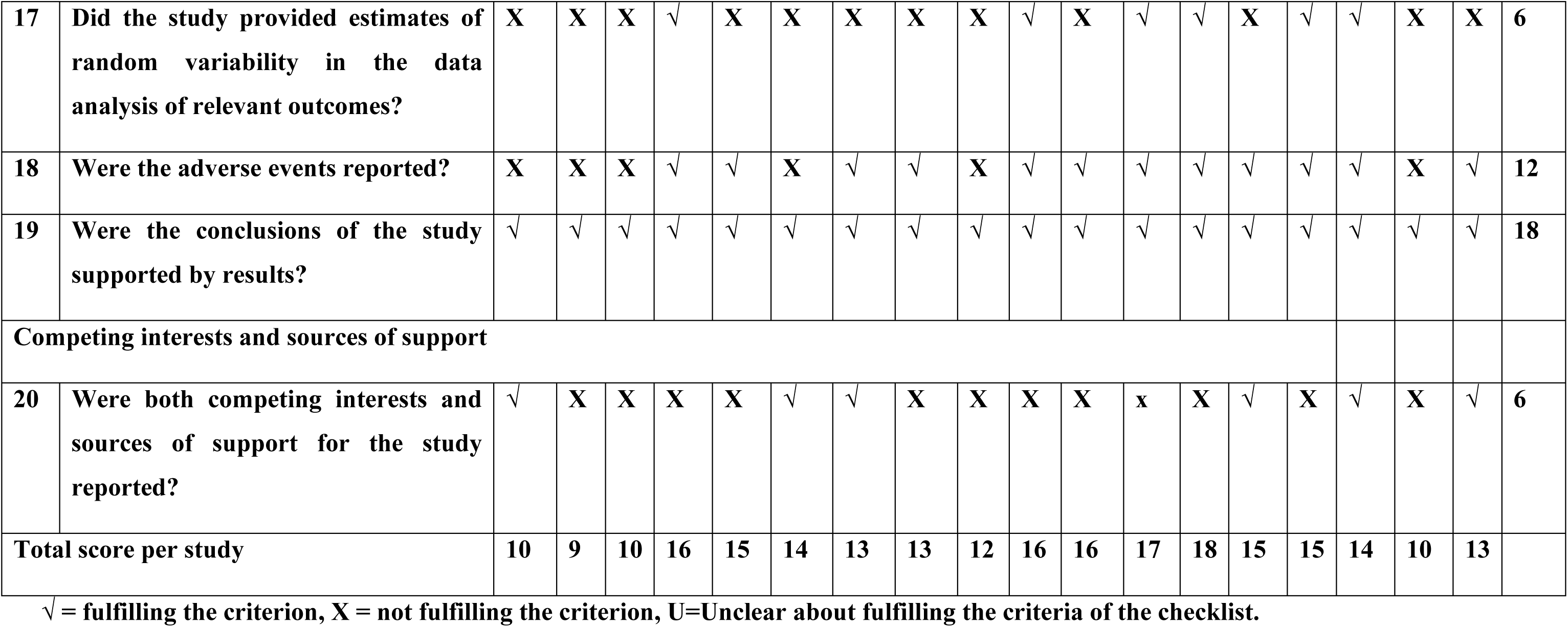
Quality assessment of the included studies as per Institute of Health Economics (IHE) checklist.

### 3.4 Meta-analysis

A total of 25 articles were contributed to qualitative synthesis and 18 (n=1212 patients) included for quantitative synthesis (meta-analysis) (Figure 1). Individual study characteristics including patient demographics, disease complications and tocilizumab dose and duration are presented in Table 1. The mortality rate of COVID-19 patients in the included case series treated with tocilizumab group was 21% (251/1212) with the pooled adjusted overall estimate 0.189 (95% CI 0.137-0.253). Across studies, there was a large positive effect of tocilizumab in reducing mortality as indicated by the forest plot. Among the included studies, 4 studies reported zero deaths in tocilizumab treated patients (Figure 2). Forest plot analysis of the primary outcome, mortality (Figure 2) also shows substantial heterogeneity among the included studies (I^2^=78.89%, p<0.001). A visual assessment of the studies’ results suggests between-study variability and the majority of individual study point estimates of the treatment effect are on the negative side of the line of overall effect but do not overlap, indicating a difference in treatment effect magnitude among studies. The confidence intervals for each study’s treatment effect (horizontal lines) do not overlap one another, and the upper and lower limits of the CI consistently line up on vertical axis, indicating a treatment effect among studies near to overall proportion effect.

**Figure 2.**
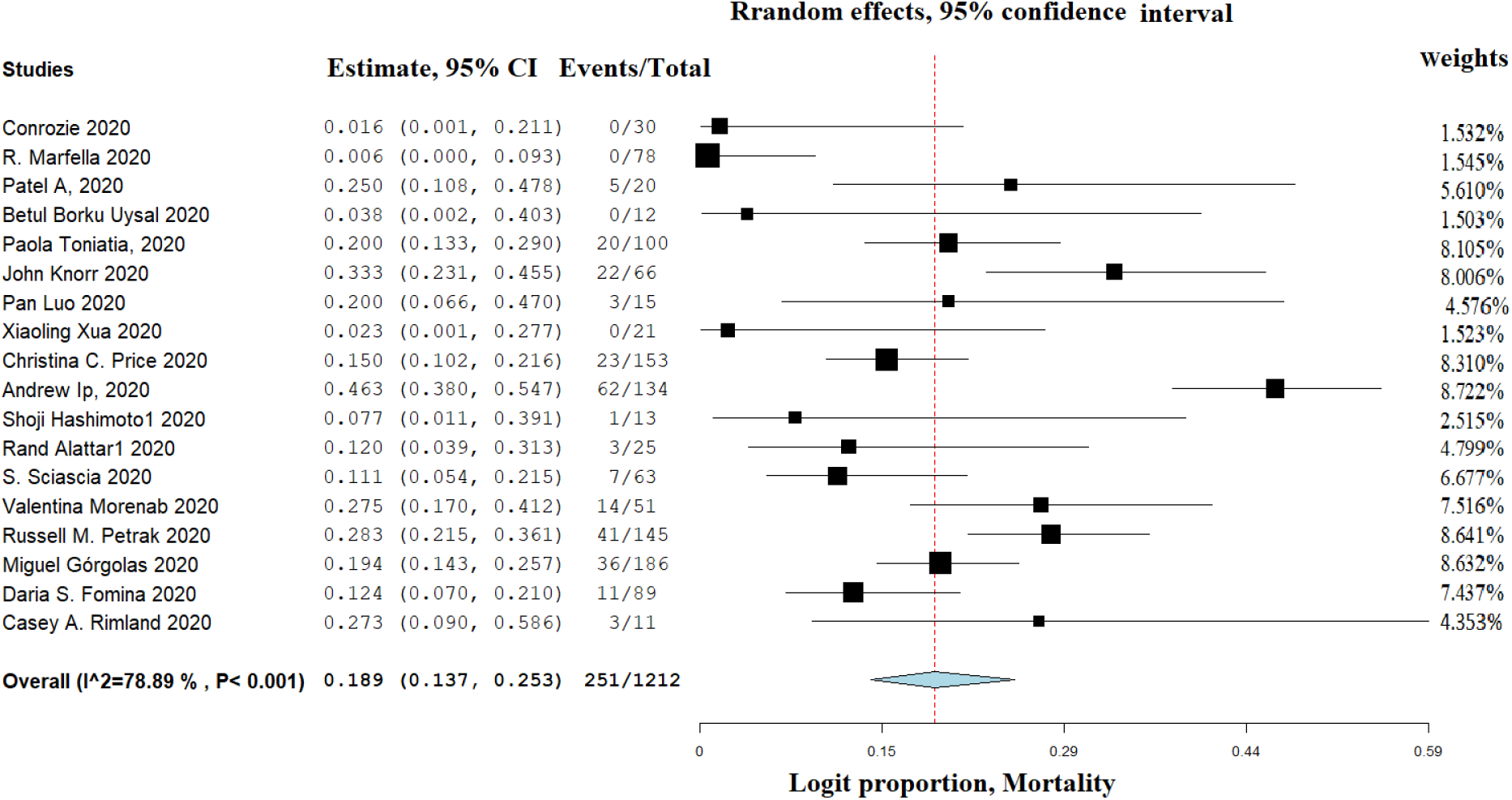
Forest plot of effect of tocilizumab on reduction in mortality of COVID 19 patients following treatment

Figure 3 and 4 represent the forest plots of effect sizes (adjusted logit proportions) for the tocilizumab effect on clinical improvement and the need for the mechanical ventilation for the patients following treatment with tocilizumab respectively. Figure 3 shows the results for 13 studies that included the data for 721 COVID 19 patients that underwent treatment with tocilizumab. The overall estimate displays a positive effect of tocilizumab with nearly 61% of patients getting improved (overall estimate and 95% CI 0.621 [0.508,0.722]. Similarly, figure 4 shows the results for 15 studies that included the data for 1091 COVID 19 patients that underwent treatment with tocilizumab. The overall estimate displays a positive effect for tocilizumab with nearly 15% of patients only needed to be mechanically ventilated (overall estimate and 95% CI 0.158 [0.107,0.227]. These forest plots, however, (Figure 3 and 4) also show substantial heterogeneity among the included studies (I^2^=83.7% and 81.3% respectively). A visual assessment of the studies’ results in these plots suggests between-study variability and more individual study point estimates of the treatment effect are on the positive side of the line of overall effect (figure 3) effect and conversely more on negative in figure 4 indicating an overall beneficial effect following treatment with tocilizumab.

**Figure 3.**
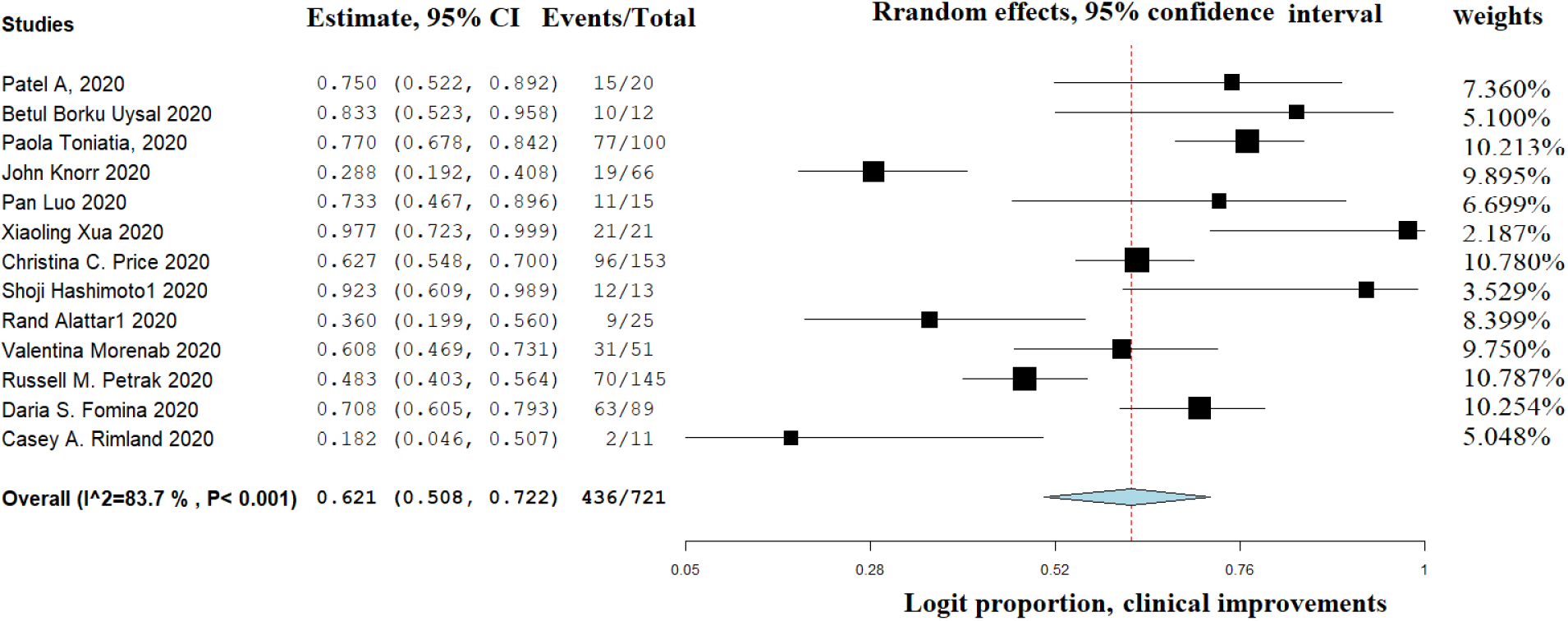
Forest plot of effect of tocilizumab on increase in clinical improvement in COVID 19 patients

**Figure 4.**
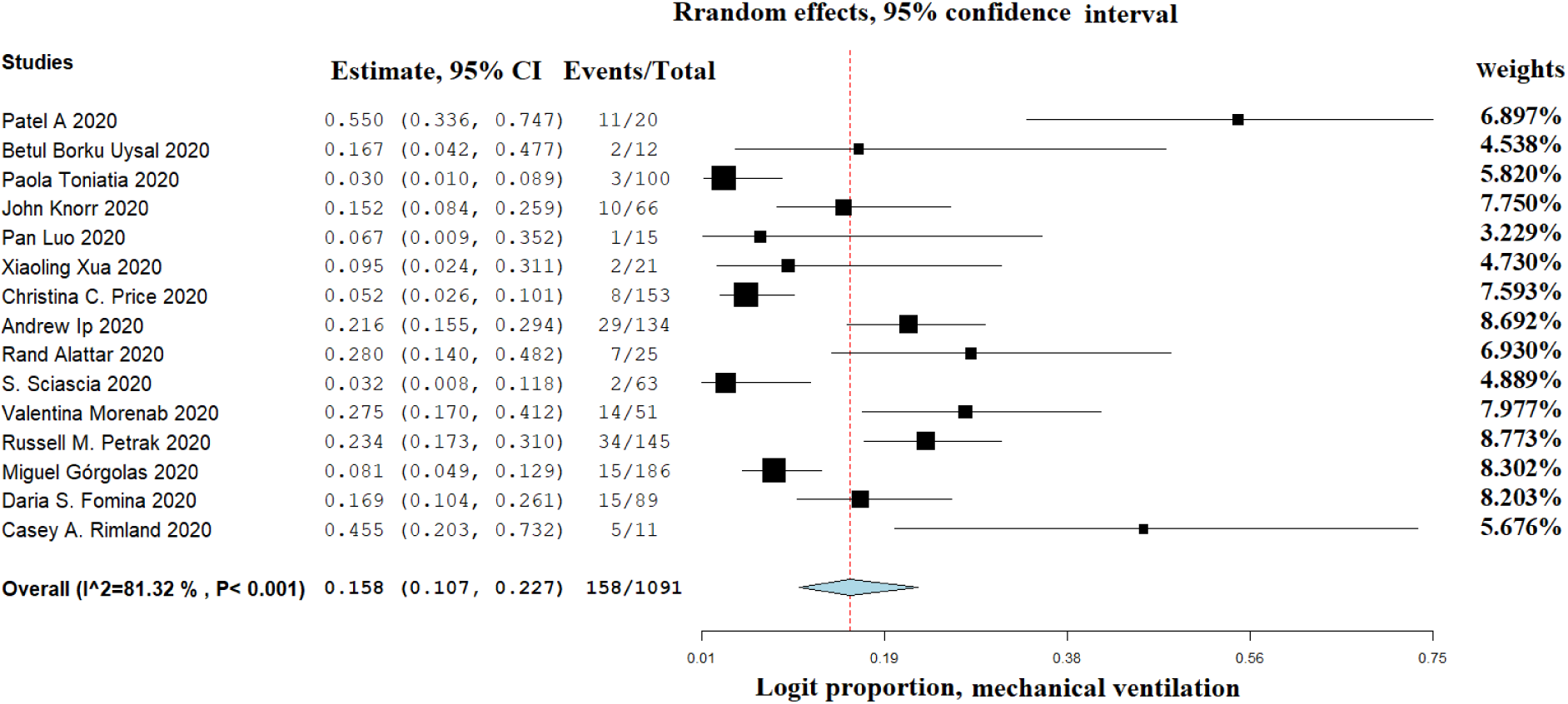
Forest plot of effect of tocilizumab on reduction in need for mechanical ventilation in COVID 19 patients

### 3.5 Assessment of publication bias of included studies

Figure 5 represents a relationship between treatment effect and study precision giving an asymmetric funnel plot in the cylindrical form due to publication bias or differences between higher and lower precision studies (typically ‘small study effects’). There was one study as an outlier falling outside of funnel. This asymmetry may also be arising due to use of an inappropriate effect measure warranting further investigation of possible causes.

**Figure 5.**
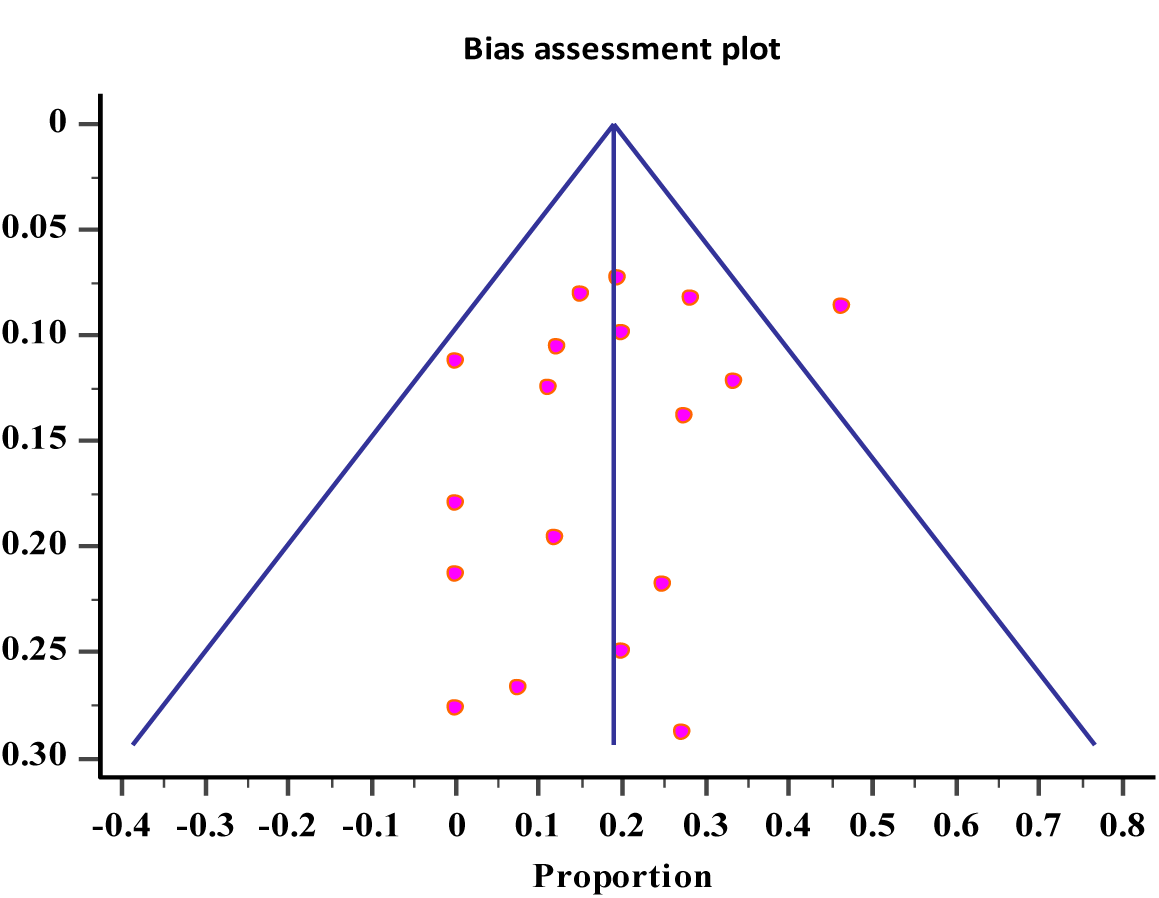
Funnel plot of case series studies regarding clinical efficacy in tocilizumab by Egger test Bias assessment plot

## 4 DISCUSSION

Tocilizumab is a humanized monoclonal antibody capable of interfering with the IL-6 binding and activation. High levels of IL-6 have been implicated in cytokine storm, cytokine release syndrome (CRS) and hypercoagulable state which are responsible for potentially causing life-threatening multiorgan damage and high risk of deaths[35]-[40]. Evidence on efficacy and safety of tocilizumab for COVID-19 from the indirect preclinical data suggests rationale for using tocilizumab and observational studies suggest that treatment with tocilizumab may be associated with more favorable outcomes compared to standard care in patients with severe or critical COVID-19[6]. Until now no RCTs have been published or made available pre-print for the effectiveness and safety of tocilizumab in the context of COVID-19. Several observational studies have examined whether tocilizumab has any effect in patients with COVID-19. Although many of the patients included in such studies had severe or critical disease and many were admitted to ICU. Drawing conclusions from such findings is a leap of faith, given that most had small sample sizes and high or moderate risk of bias.(37-40)

The assessment of quality of publications had score from 9 to 15 out of a possible 20 points indicating the studies to be of poor to moderate quality according to the “Quality Appraisal Checklist for Case Studies” tool developed by the Institute of Health Economics[11].

There was a large variation in the characteristics of selected studies and do not seem to have satisfactory follow up duration and majority of them were retrospective in nature. Eighteen studies assessed the mortality of COVID 19 patients following treatment with tocilizumab with an overall mortality rate 21% (251/1212)). The other metanalysis carried to assess efficacy of tocilizumab in case controlled and cohort studies also reported similar mortality rate of 22.4% (258/1,153) when compared to control[6]. However it is noteworthy to mention there was a substantial heterogeneity (I^2^=78.9%) in our study which is also similar to other published metanalysis (I^2^=80%) [6]. The possible reasons for heterogeneity being, difference in the age, study design, treatment strategies, presence of comorbidities and variability in the follow-up period. About thirteen studies also reported clinical improvements in COVID 19 patients (436/721) and fifteen studies (158/1091) assessed the need for mechanical ventilation by the patients.[9], [16], [24]–[32], [17]–[20], [20]–[23] [9], [16], [25], [27], [29]–[31],[34], [17]–[24] Majority of the case series reported were found to from USA- and Italy. Common co-morbidities reported in COVID 19 patients were hypertension, diabetes mellitus, chronic pulmonary disorder, obesity, rheumatoid arthritis and kidney disease. Only few studies treating COVID 19 patients with tocilizumab did not report any adverse events. Majority of the studies adopted standard of care treatment (SOC) along with one to three doses of tocilizumab. The results generated in the current meta-analysis should be taken in context with the evidence available for the efficacy of tocilizumab in the treatment of COVID 19 infections in the best interest of tackling this pandemic. As case reports and case series are uncontrolled study designs with known risk of bias but should help us in this tiring time to advance our knowledge and preparedness.

**In conclusion** the present synthesis and inference derived from case reports and case series should provide us useful insights with the other available evidence to refine our strategy and equip ourselves effectively with tocilizumab to defeat COVID 19 to save humanity. We suggest using evidence derived from the metanalysis of the current case series to inform decision-making about the efficacy of tocilizumab for COVID 19 until the higher level of evidence is available through randomized controlled trials.

### Limitations

The included studies utilized varied doses of tocilizumab (single or double), and duration drug availability issues emerged in some centers, which may have influenced both sample sizes and study designs.

The treatment allocations in these observational studies were based solely upon physician judgement rather than random assignment thereby leading to chances of risk of bias without accounting for risk factors. As the case series consists of studies with low sample sizes might likely cause over estimation of the overall effect size.

## Data Availability

NONE

## Clinical implications

Incorporating studies without control groups into systematic reviews and quantitative synthesis especially when there are no other studies to consider can provide information for formulating effective treatment strategies for management of COVID 19 infections through the use of tocilizumab.

Clinicians should continue compassionate use of tocilizumab as an option for COVID-19 patients and enroll COVID-19 patients in clinical trials for assessing the safety and efficacy of tocilizumab until the future RCTs enlighten the clinical efficacy of tocilizumab.

